# Economic Burden of Measles among Hospitalised Children in Kenya

**DOI:** 10.64898/2026.01.23.26344606

**Authors:** Prashant Mandaliya, Stacey Orangi, Isaac Waluke, Franklin Okech, Felix Masiye, Obinna Onwujekwe, Edwine Barasa

**Affiliations:** Health Economics Research Unit, KEMRI-Wellcome Trust Research Programme, Nairobi, Kenya; Health Services Unit, KEMRI-Wellcome Trust Research Programme, Nairobi County, Kenya; Department of Economics, School of Humanities and Social Sciences, University of Zambia, Zambia; University of Nigeria, Nsukka, Nigeria; Centre for Global Health and Tropical Medicine, Nuffield Department of Medicine, University of Oxford, Roosevelt Drive, Oxford OX3 7LG, UK

**Keywords:** measles, paediatric, cost-of-illness, hospitalization, catastrophic health expenditure, health economics, health financing

## Abstract

Measles is endemic in Kenya, mainly affecting young children, with low vaccination coverage leading to recurrent outbreaks. No published study has estimated the cost of treating hospitalised paediatric measles patients in Kenya. This study quantified the cost of treating measles from the healthcare provider and societal perspectives, identified the main cost drivers, and assessed the proportion of households likely to experience catastrophic health expenditure due to the disease.

A retrospective, prevalence-based cost-of-illness study was conducted using data from the Clinical Information Network (CIN) for 214 children hospitalised with measles across 16 Kenyan public hospitals from 2013 to 2024. Quantities of resources were extracted from CIN data, and unit costs were obtained from price lists, facility surveys, expert interviews, and market surveys. All costs were converted to 2025 KES and USD. A simulation-based catastrophic health expenditure (CHE) analysis was conducted. One-way sensitivity analysis varied hospital bed, staff, direct non-medical and indirect costs.

Median cost per admission was USD 139.11 [IQR: 91.58 to 186.05] (provider) and USD 176.18 [IQR: 116.68 to 256.07] (societal). Staff costs were the primary driver across both perspectives (67% provider, 53% societal), followed by bed days (21% provider, 17% societal). Adjusting the staff time increased treatment costs by 83% (provider) and 73% (societal), while varying bed charges had a lesser effect (-10% to 6% for provider and -3% to 10% for societal). CHE analysis showed that more than half of the households could face catastrophic cost if full treatment costs were paid out-of-pocket.

The cost of treating hospitalised paediatric measles patients in Kenya is significant, driven by staff and hospital-bed costs. Policymakers should utilize the evidence generated to expand insurance coverage and refine sub-national resource allocation, to help reduce the catastrophic costs on households and burden on the healthcare system.

## Introduction

While safe and effective vaccines against measles exist, the disease still remains a significant public health issue. The *Morbillivirus* causes the diseases, were infected patients typically present with fever, cough, runny nose, and a distinct maculopapular rash [1]. Global data from 2023 indicates that more than 10 million cases and over 100,000 deaths occurred from measles in that year [2]. The disease burden is greater in younger children [2], and the prevalence remains more pronounced in low- and middle-income countries (LMICs) [3]. In Kenya, the disease is endemic and a recurrent public health concern [4], with the number of cases between 2003 and 2016, exceeding 9,000 [5]. A recently published WHO article reported 525 measles cases in Kenya for the year 2024 [6].

For countries to reach optimal protection, WHO recommends vaccine coverage of 95% for both doses of the vaccine [7]. As of 2024, the official vaccine coverage in Kenya for the first dose of measles-containing vaccine (MCV1) was 82%, with the second dose (MCV2) being far lower at 62% [8]. The low vaccine coverage of both doses in the country has contributed to the recurrent outbreaks seen in the country [5]. Given these challenges, understanding the cost of treating the disease will provide evidence on the economic burden of measles.

Cost-of-illness (COI) studies help quantify the costs incurred during the treatment of a disease [9]. These studies can include the direct medical costs (expenditures on healthcare services), direct non-medical costs (transport, food, accommodation), and indirect costs (lost productivity due to illness or caregiving), depending on the perspective employed [10]. A recently published systematic review highlighted that treatment costs for measles are highly contextual, and also underscored the lack of published literature from LMICs [11]. Treatment of measles can impose severe financial costs on African households[12,13], leading to catastrophic health expenditure (CHE). CHE is defined as the costs related to healthcare that exceeds a household’s capacity to pay [14,15]. As per standard WHO methodology, capacity to pay (CTP) is typically defined as 40% of the annual non-food household expenditure [16]. Evidence on CHE can help policymakers understand the level of out-of-pocket (OOP) spending households of different wealth levels can bear before facing impoverishment.

This study provides the first estimates for the cost of treating hospitalised paediatric measles patients in Kenya, an important contribution given the endemicity of the disease in the country. Furthermore, it estimated the proportion of households that could face CHE under different scenarios due to the disease. The evidence generated by the study would help policy makers better understand the financial impact of the disease on households and the healthcare system. It would also provide a justification for prioritising investments to improve vaccination and national health insurance coverage. Researchers conducting economic evaluations of alternative measles vaccination strategies for Kenya or other sub-Saharan countries, can utilize the cost estimates reported in the study as model inputs.

## Methodology

### Study setting

Kenya is a LMIC with nearly half of the estimated 47.5 million people (as of 2019), being 18 years or younger [18]. The country’s gross domestic product (GDP) per capita in 2024 was United States Dollars (USD) 2206 [19]. Kenya is devolved into 47 counties that are semi-autonomous and responsible for delivering public health services [20]. Most of the healthcare-related functions are managed at the county level (70%), while the national government handles the remaining responsibilities [21]. Healthcare facilities that are county managed receive funding through the county health budget, which are supplemented by reimbursements from the public insurer and OOP payments made by patients [22].

### Study design and study population

The study utilised a retrospective, prevalence-based cost-of-illness approach to evaluate the economic burden of measles amongst children under 15 years in Kenya and to assess the incidence of CHE using a simulation-based methodology [23]. Patient-level data was obtained from the Clinical Information Network (CIN), which routinely collects admission, care and discharge data of neo-nates and paediatric patients across 25 public hospitals, covering close to half of the counties (19 out of 47) in the country [24].

### Costing approach, perspective, and time horizon

Our analysis followed a bottom-up micro-costing approach, which involved identifying, measuring, and valuing individual resources used in managing measles cases [25]. The perspectives employed included the healthcare provider and societal perspectives, with a time horizon limited to the period a patient was hospitalization.

### Cost categories

Full economic costs were estimated by including the monetary value of all inputs, whether financial payments were involved or not. Costs were classified as direct medical costs, direct non-medical costs, and indirect costs, which are detailed in Figure 1.

**Figure 1:**
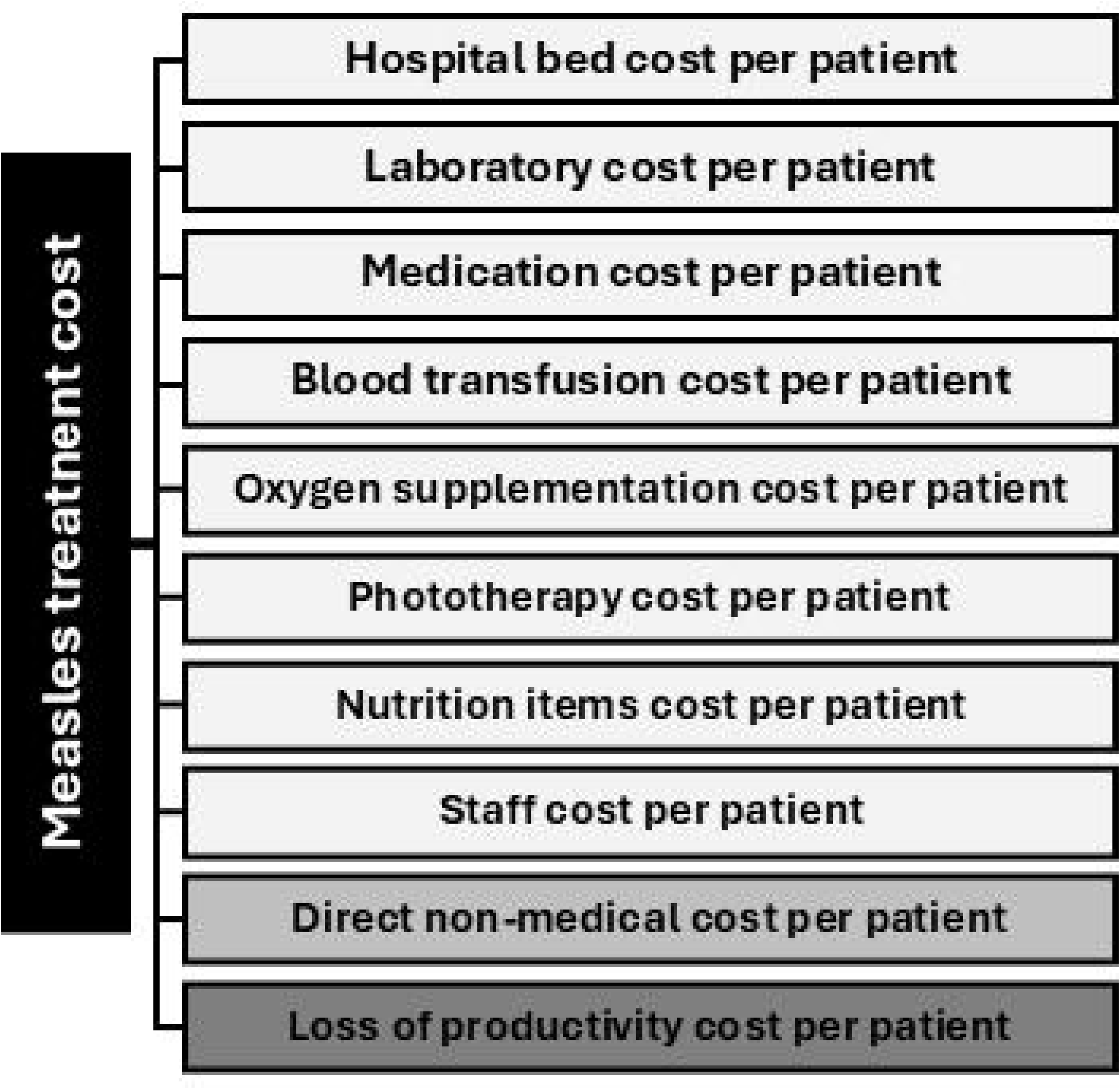
Cost categories for the treatment of measles.

From the healthcare provider perspective, only direct medical costs were considered, whereas the societal perspective incorporated direct medical, direct non-medical, and indirect costs to provide a more comprehensive estimate of the economic burden.

### Data collection

Patient-level data was sourced from the CIN database, which is stored on REDCap 13.1.5 [26,27]. This database includes over 600 clinical and demographic variables, offering a comprehensive details of the included patients [24]. The CIN team identified and extracted records of patients diagnosed with measles over a 11 year period (2013 to 2024), using R 4.1.2 [28]

The extracted data, underwent steps for data cleaning and handling of missing values, which were conducted using STATA v17 [29]. The data cleaning process involved removal of duplicate records, standardization of diagnostic codes, and resolving treatment related inconsistencies. Missing data were addressed through imputation methods, that utilised the mean, mode, and literature-based assumptions. All assumptions for missing data were first validated through consultations with experts (specialists, medical officers, pharmacists, and nurses) to ensure that they were applicable in the Kenyan context.

### Measuring and valuing resources

The first step involved identifying and quantifying the resources used by the patients and assigning unit costs to each cost item. The quantities for each item were obtained from the CIN data and expert opinion, whereas unit costs were collected from relevant price lists and surveys (detailed in subsequent sub-sections). Finally, the total cost for each item (*i*) was calculated:

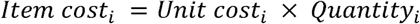

Items belonging to the same cost category (*k*) were then summed to obtain the total cost for each cost category:

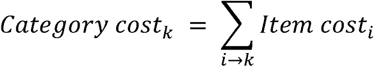

All unit costs were first converted to the local currency unit which was Kenya shillings (KES) based on the average annual exchange rate for the base year and then inflated to 2025 KES using GDP deflators for Kenya [30,31]. For the years 2024 and 2025, for which official GDP deflators were unavailable at the time of the analysis, an annual inflation rate of 5.3% was used to determine the deflator values [32]. All cost estimates reported in 2025 KES were subsequently converted to USD using an exchange rate of KES 129.28 per USD [33]. The process of measuring and valuing each cost category is detailed in the subsequent sub-sections.

### Direct medical costs

#### **i)** Hospital bed costs

The number of days each patient was hospitalised for, was derived from the admission and discharge dates reported in the CIN dataset. Daily bed costs covering accommodation and overheads were obtained through a survey of 14 public hospitals (Table S1 in Appendix 1). The median value from the survey was utilised to represent the daily hospital bed cost for all patients.

#### **ii)** Pharmaceuticals and non-pharmaceuticals costs

The quantity of each pharmaceutical consumed during treatment was determined by examining the prescription-related variables in the CIN dataset. This included the drug name, route, strength, frequency, and duration of treatment. The total amount of pharmaceuticals consumed by each patient was calculated as:

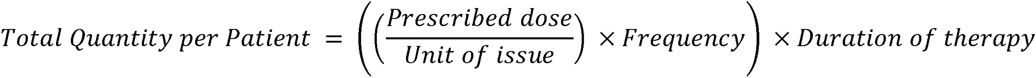

We determined the quantities for non-pharmaceuticals and nutritional support items by aggregating the number of times each item was prescribed.

The unit costs of pharmaceuticals, non-pharmaceuticals, and nutritional items were obtained using the approach generally followed by the procurement department at public hospitals in Kenya [34]. The first data source was the pricelist for Kenya Medical Supplies Authority (KEMSA), chosen because of the company’s role as the primary supplier of medical items to public health facilities in Kenya [35]. For items that were unavailable at KEMSA, the pricelist for the Mission for Essential Drugs and Supplies pricelist was consulted [36], with any items missing from these sources were obtained through a market survey of local suppliers in Nairobi, using convenience sampling [37].

#### **iii)** Laboratory, radiology, blood transfusion, phototherapy and oxygen costs

Total quantities for the different laboratory tests and radiological procedures were obtained by determining the number of times each were requested and then summing the individual requests to obtain the total quantity. The same process was followed to determine total quantities for blood transfusions, phototherapy sessions, and oxygen supplementation.

Unit costs for laboratory and radiology services were taken from a 2023 cost survey of seven public hospitals in Kenya [38], with the median values used to represent the unit cost of each service. For tuberculosis-related tests, which are offered free in public facilities, private sector costs were used [39]. Details are provided in Table S2 under Appendix 2.

The unit cost of blood transfusion was obtained from a survey of three public hospitals, with the median cost of USD 7.74 utilised (see Table S3 in Appendix 3). Cost for oxygen supplementation (USD 16.81) was sourced from Kenya-specific literature [40], while phototherapy cost per session (USD 2.94) was obtained from Kenyatta National Hospital, the country’s largest public referral hospital [41].

#### **iv)** Staff costs

To estimate the quantity of time spent by staff to care for measles patients, health workers of different cadres (paediatricians, medical officers, clinical officers, nurses, and nutritionists) working in public hospitals, were consulted for their expert opinions. Respondents provided estimates of the time they would spend to care for a patient with measles in a public hospital over a 24-hour period, on routine care tasks such as assessment, monitoring, prescribing, administering medications, counselling, and documentation. Purposive sampling was employed until saturation was reached [42]. The mean time for each cadre was applied as the quantity (Table S4 in Appendix 4).

Unit costs for staff time were calculated using data on monthly salaries, which were sourced from a study by Barasa et al. [40]. The study reported the official salaries of healthcare worker of different cadres working in the Kenyan public health sector. Monthly salaries were converted to per-minute values, with the assumption that a healthcare worker would work 40 hours per week in accordance with Kenyan human resource policies [43].

### Direct non-medical cost

To determine the direct non-medical expenditure, unit costs were adapted from a recent study looking at the cost of treating acute illnesses in hospitalised children in Kenya and Uganda [44]. The reported values were converted to per-day direct non-medical costs (disaggregated by geographical setting), using the weighted average cost and weighted average length of stay for urban and rural counties. The unit cost estimates were then adjusted to 2025 KES and converted to USD (Urban: USD 4.59; Rural: USD 2.18). The length of each patient’s hospital stay was used as the measure of quantity.

### Indirect costs

The human capital approach was used to estimate the indirect costs incurred. The daily wage rates for general labourers (minimum wage) in Kenya were used as the unit costs [45]. These costs were disaggregated by county groups. The duration of hospitalization represented the quantity.

### Cost function

The total cost of illness was calculated by summing the cost categories for each patient, based on the perspective employed:

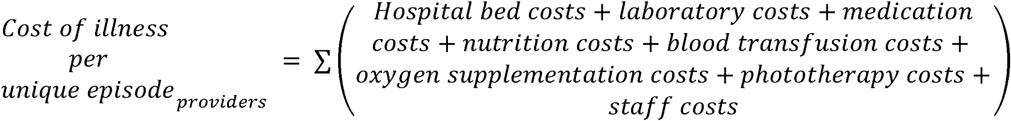

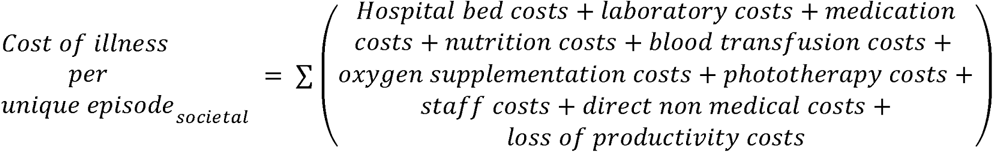

### Sub-group analysis

The mean treatment cost and mean cost for each cost category was estimated using a generalised linear model (GLM) with a gamma distribution and log-link function. The model type was appropriate for the right skewed distribution of cost data [46]. The outputs of the GLM included the mean estimates with 95% confidence intervals (CIs).

The sub-group analyses looked at how the cost varied by gender, age-group, geographical setting, hospital level, referral status, diagnosis, vaccination status and county. The percentage difference in mean costs across each group were estimated using GLM, with statistical significance (p<0.05) evaluated for inter group differences between the sub-group(s) and reference group (see Table S5 in Appendix 5 for details on the selected reference categories). A similar approach was adapted for median values; were the Wilcoxon rank-sum test was used to determine if the differences were statistically significant. All analyses were performed using STATA v17.

### Sensitivity analysis

To test the robustness of our cost estimates, we carried out a one-way sensitivity analysis focusing on four cost categories: hospital bed day costs, staff costs, direct non-medical costs and loss of productivity costs. Each of the selected parameters was varied independently to see how changes would affect the total cost of illness from both the healthcare provider and societal perspectives, while holding all other parameters constant.

For hospital bed day costs, the 25th and 75th percentile values represented the lower and upper limits, respectively. Since time-use data specific to measles care was obtained from expert opinion, we adjusted the reported values using a factor derived from the Harmonised Health Facility Survey 2018–2019 [47], to test the robustness of the estimates. Derivation of the adjustment factor and calculation of the adjusted staff times are detailed in Table S6 under Appendix 6.

The two cost parameters adopted from literature were also included in the sensitivity analysis. For direct non-medical costs, the values were varied between the lower and upper CIs, while productivity loss costs were varied by ±20 percent from the base value to obtain the upper and lower limit.

### Catastrophic health expenditure

We estimated the incidence of CHE among households of measles patients using a simulation-based approach proposed by Mandaliya et al. [23]. The detailed methodology is found in Appendix 7, Table S7 and Table S8. The proposed methodology is a modified version of that described by Xu et al. [16], and was used as no OOP payment and household level expenditure data was collected from patients.

The total cost a patient could pay OOP was determined by summing the direct medical and non-medical costs. The total cost paid by each patient was then varied across six different levels of OOP expenditure (20%, 24.2%, 40%, 60%, 80% and 100%). The base value of 24.2% represented the average OOP health expenditure in Kenya [48]. The non-food household expenditure (NFHE) for each wealth quintile, covering both urban and rural households, were adopted from the literature [23]. The authors calculated the NFHEs using data obtained from national surveys conducted in Kenya [18,49–52].

A patient’s household experienced CHE, if the CHE ratio (R) of OOP to NFHE was greater than 0.4 [16]. The R was computed as:

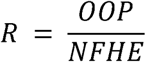

Each household was simulated across thirty unique scenarios to determine under which scenario would the household face CHE. A patient’s household was assigned 1 if it experienced CHE, else 0. These values were then summed across each scenario to determine the number of households that could face CHE in the given scenario. The reported outcomes were disaggregated by geographical setting.

## Results

### Patient demographics

A total of 214 children diagnosed with measles from 16 of the 42 counties were included in the analysis as detailed in Table 1. Most patients were male (63%) with a median age of 16 months [IQR: 9 - 42 months]. Patients were mainly direct admissions (90%), with a larger proportion coming from hospitals in rural areas (65%). Less than 20% of the patients were recorded to have taken at least one dose of the measles vaccine. The median duration of hospitalization was 6 days.

**Table 1.** Demographic and referral characteristics of the study population.

| Name | Patient count | Percent (%) |
| --- | --- | --- |
| **Gender** |  |  |
| Male | 135 | 63.1 |
| Female | 79 | 39.9 |
| **Age-group** |  |  |
| Under 1 year | 74 | 34.6 |
| 1-4 years | 110 | 51.4 |
| 5 years and older | 30 | 14.0 |
| **Setting type** |  |  |
| Rural | 138 | 64.5 |
| Urban | 76 | 35.5 |
| **Hospital county** |  |  |
| Bungoma | 1 | 0.5 |
| Busia | 18 | 8.4 |
| Embu | 7 | 3.3 |
| Homabay | 4 | 1.9 |
| Kakamega | 4 | 1.9 |
| Kiambu | 20 | 9.4 |
| Kilifi | 47 | 22.0 |
| Kirinyaga | 4 | 1.9 |
| Kisumu | 14 | 6.5 |
| Machakos | 19 | 8.9 |
| Migori | 2 | 0.9 |
| Murang'a | 3 | 1.4 |
| Nairobi | 52 | 24.3 |
| Nakuru | 4 | 1.9 |
| Nyeri | 10 | 4.7 |
| Trans Nzoia | 5 | 2.3 |
| **Hospital level** |  |  |
| 4 | 70 | 32.7 |
| 5 | 143 | 66.8 |
| 6 | 1 | 0.5 |
| **Referral status** |  |  |
| Referred to the hospital | 23 | 10.8 |
| Not referred to the hospital | 191 | 89.3 |
| **Vaccination status** |  |  |
| Vaccinated | 40 | 18.7 |
| Unvaccinated | 174 | 81.3 |

### Cost of treatment

The median total cost of treatment pre unique episode from the healthcare provider perspective was USD 139.11 (mean: USD 155.06). Staff costs accounted for the largest proportion of the median treatment costs (67%), with hospital bed costs also being significant (21%). Services which were not routinely used, such as blood transfusion, oxygen supplementation, phototherapy and nutrition items were grouped together (other costs) and accounted for less than 1% of the treatment costs. This information is further detailed in Table 2.

**Table 2:**
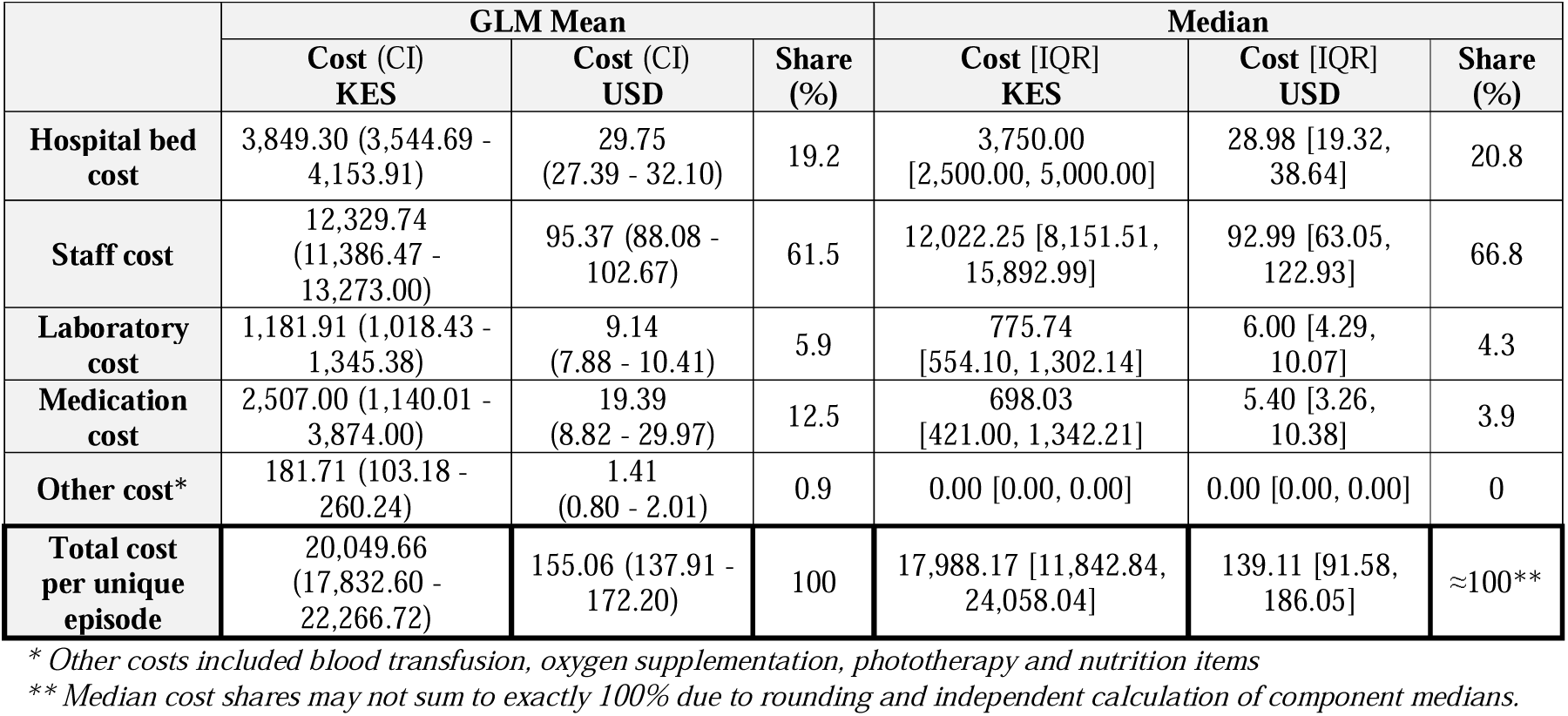
Median cost of treatment from the healthcare provider’s perspective.

From the societal perspective, the median cost of treatment was about 27% higher at USD 176.18 (mean: USD 205.11). Staff and hospital bed costs remained the largest contributors to total treatment costs, with productivity loss and direct non-medical costs accounting for 14% and 8% respectively, as detailed in Table 3.

**Table 3:**
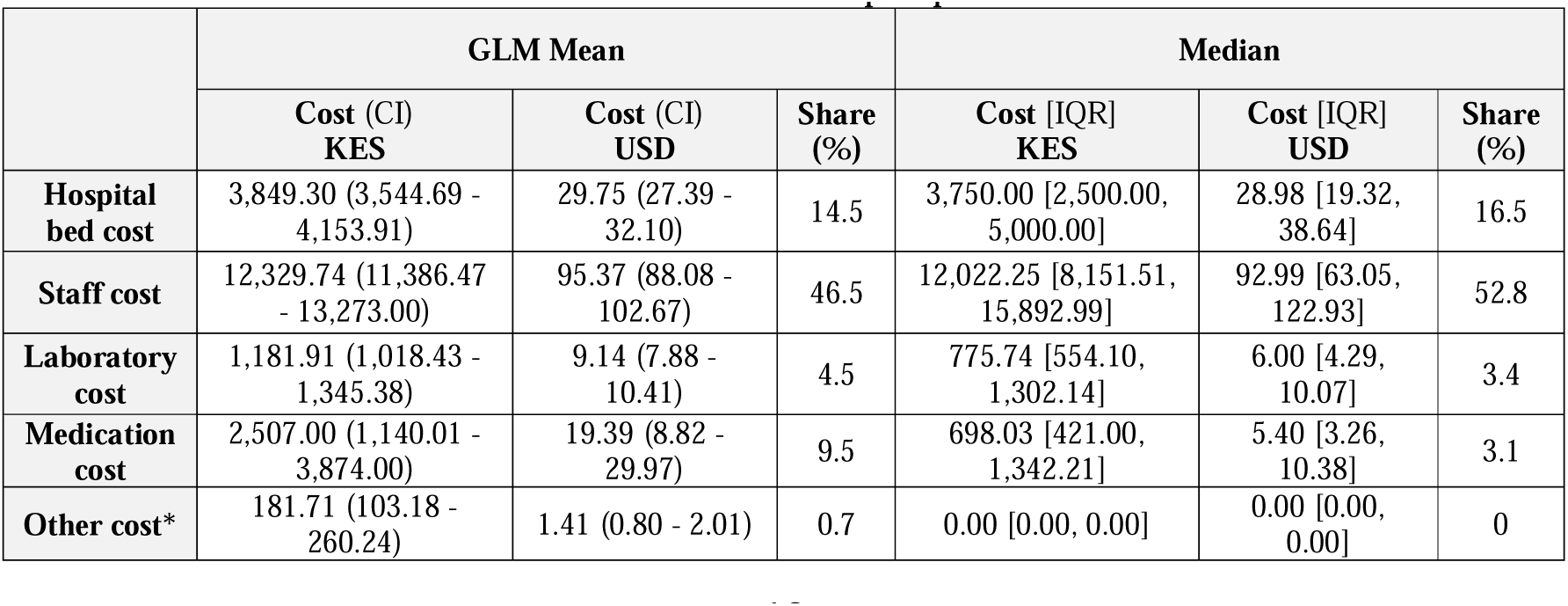
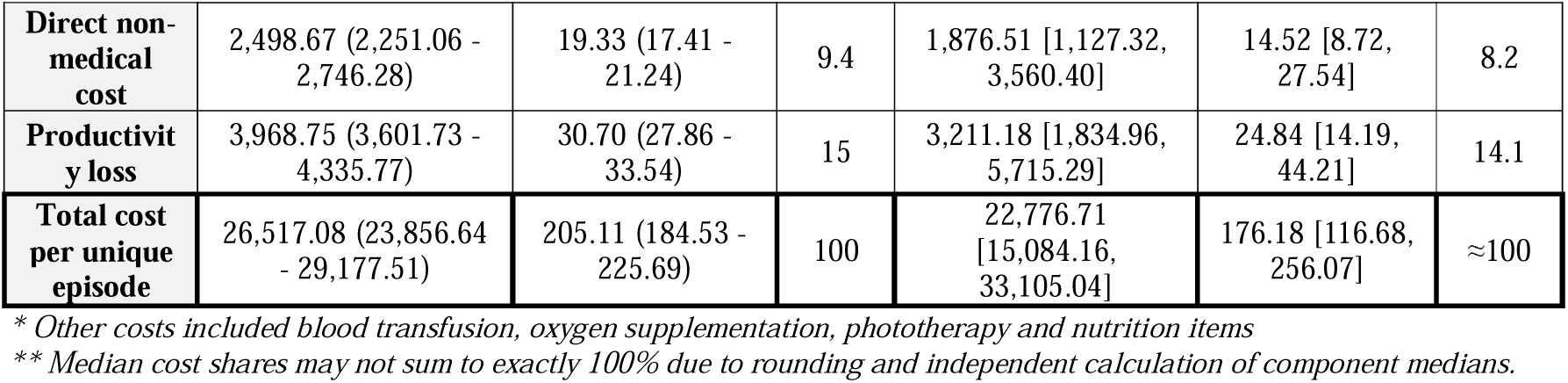
Median cost of treatment from the societal perspective.

|  | GLM Mean |  |  | Median |  |  |
| --- | --- | --- | --- | --- | --- | --- |
|  | Cost (CI)<br>KES | Cost (CI)<br>USD | Share<br>(%) | Cost [IQR]<br>KES | Cost [IQR]<br>USD | Share<br>(%) |
| **Hospital bed cost** | 3,849.30 (3,544.69 - 4,153.91) | 29.75 (27.39 - 32.10) | 14.5 | 3,750.00 [2,500.00, 5,000.00] | 28.98 [19.32, 38.64] | 16.5 |
| **Staff cost** | 12,329.74 (11,386.47 - 13,273.00) | 95.37 (88.08 - 102.67) | 46.5 | 12,022.25 [8,151.51, 15,892.99] | 92.99 [63.05, 122.93] | 52.8 |
| **Laboratory cost** | 1,181.91 (1,018.43 - 1,345.38) | 9.14 (7.88 - 10.41) | 4.5 | 775.74 [554.10, 1,302.14] | 6.00 [4.29, 10.07] | 3.4 |
| **Medication cost** | 2,507.00 (1,140.01 - 3,874.00) | 19.39 (8.82 - 29.97) | 9.5 | 698.03 [421.00, 1,342.21] | 5.40 [3.26, 10.38] | 3.1 |
| **Other cost\*** | 181.71 (103.18 - 260.24) | 1.41 (0.80 - 2.01) | 0.7 | 0.00 [0.00, 0.00] | 0.00 [0.00, 0.00] | 0 |
| **Direct non-medical cost** | 2,498.67 (2,251.06 - 2,746.28) | 19.33 (17.41 - 21.24) | 9.4 | 1,876.51 [1,127.32, 3,560.40] | 14.52 [8.72, 27.54] | 8.2 |
| **Productivity loss** | 3,968.75 (3,601.73 - 4,335.77) | 30.70 (27.86 - 33.54) | 15 | 3,211.18 [1,834.96, 5,715.29] | 24.84 [14.19, 44.21] | 14.1 |
| **Total cost per unique episode** | 26,517.08 (23,856.64 - 29,177.51) | 205.11 (184.53 - 225.69) | 100 | 22,776.71 [15,084.16, 33,105.04] | 176.18 [116.68, 256.07] | ≈100 |

\* Other costs included blood transfusion, oxygen supplementation, phototherapy and nutrition items
\*\* Median cost shares may not sum to exactly 100% due to rounding and independent calculation of component medians.

### Subgroup analysis

The differences in the cost of treatment for measles, from the societal perspectives across various sub-groups are shown in Table 4. The findings from the healthcare provider’s perspective are presented in Table S9 under Appendix 8. Majority of the sub-groups showed differences that were statistically significant, with gender and vaccination status being the only ones that did not have significant difference within the subgroup.

**Table 4:** Median costs for treatment of measles by subgroup.

| Name | N (%) | Mean total cost (95% CI) |  |  | Median total cost [IQR] |  |  |
| --- | --- | --- | --- | --- | --- | --- | --- |
|  |  | 2025, USD | % Difference (CI) | P-value | 2025, USD | % Difference (CI) | P-value |
| Gender |  |  |  |  |  |  |  |
| Female | 79 (36.9) | 219.48 (182.79 – 256.17) | Reference | - | 182.91 [127.31 – 278.53] | Reference | - |
| Male | 135 (63.1) | 196.71 (171.55 – 221.86) | -10.4% (-27.4% to 10.6%) | 0.308 | 171.17 [113.60 – 241.57] | -6.4% (-26.3% to 13.4%) | 0.107 |
| Age-Group |  |  |  |  |  |  |  |
| Under 1 year | 74 (34.6) | 212.97 (177.15 – 248.80) | Reference | - | 198.80 [131.41 – 276.64] | Reference | - |
| 1-4 years | 110 (51.4) | 211.12 (181.99 – 240.25) | -0.9% (-20.3% to 23.2%) | 0.937 | 173.77 [113.79 – 241.57] | -12.6% (-31.9% to 6.7%) | 0.114 |
| 5 years and older | 30 (14.0) | 163.70 (120.45 – 206.95) | -23.1% (-43.8% to 5.1%) | 0.1 | 146.93 [92.85 – 229.09] | -26.1% (-53.9% to 1.7%) | 0.018* |
| Geographical setting |  |  |  |  |  |  |  |
| Rural | 138 (64.5) | 184.37 (160.59 – 208.15) | Reference | - | 159.80 [110.34 – 217.81] | Reference | - |
| Urban | 76 (35.5) | 242.78 (200.58 – 284.98) | 31.7% (6.1% to 63.5%) | 0.013* | 229.21 [157.70 – 285.50] | 43.4% (17.2% to 69.7%) | 0.001* |
| Hospital level |  |  |  |  |  |  |  |
| Level 5 | 143 (66.8) | 217.49 (191.85 – 243.14) | Reference | - | 198.74 [121.76 – 270.00] | Reference | - |
| Level 4 | 70 (32.7) | 174.26 (144.89 – 203.62) | -19.9% (-34.8% to -1.6%) | 0.035* | 159.80 [113.79 – 203.99] | -19.6% (-35.7% to -3.5%) | 0.027* |
| Level 6 | 1 (0.5) | 594.82 (0.00 – 1433.47) | 173.5% (-33.6% to 1025.6%) | 0.163 | 594.82 [594.82 – 594.82] | 199.30% | - |
| **Referral status** |  |  |  |  |  |  |  |
| Not referred | 191 (89.3) | 197.85 (176.45 – 219.25) | Reference | - | 169.98 [115.44 – 241.57] | Reference | - |
| Referred | 23 (10.8) | 265.45 (182.70 – 348.19) | 34.2% (-3.5% to 86.6%) | 0.081 | 260.73 [197.79 – 341.85] | 53.4% (11.3% to 95.5%) | 0.001* |
| **Diagnosis** |  |  |  |  |  |  |  |
| Measles Only | 27 (12.6) | 125.82 (92.61 – 159.03) | Reference | - | 117.60 [74.46 – 161.69] | Reference | - |
| Measles +1 | 45 (21.0) | 192.02 (152.76 – 231.29) | 52.6% (9.3% to 113.1%) | 0.013* | 170.71 [116.34 – 260.64] | 45.2% (-10.6% to 100.9%) | 0.008* |
| Measles +2 or more | 142 (66.4) | 224.34 (198.52 – 250.16) | 78.3% (33.7% to 137.8%) | 0 | 193.93 [131.41 – 273.04] | 64.9% (26.2% to 103.6%) | <0.001* |
| **Vaccination status** |  |  |  |  |  |  |  |
| Vaccinated | 40 (18.7) | 182.94 (140.73 – 225.15) | -13.0% (-32.6% to 12.4%) | 0.287 | 148.13 [99.02 – 212.90] | -18.9% (-37.6% to -0.3%) | 0.099 |
| Not Vaccinated | 174 (81.3) | 210.21 (186.95 – 233.47) | Reference | - | 182.73 [117.60 – 260.73] | Reference | - |
| **County** |  |  |  |  |  |  |  |
| Nairobi | 52 (24.3) | 263.38 (218.34 – 308.41) | Reference | - | 230.06 [185.08 – 294.25] | Reference | - |
| Bungoma | 1 (0.5) | 196.94 (0.00 – 439.78) | -25.2% (-78.5% to 159.7%)* | 0.647 | 196.94 [196.94 – 196.94] | -14.40% | -** |
| Busia | 18 (8.4) | 143.85 (102.04 – 185.66) | -45.4% (-61.0% to -23.5%) | <0.001* | 117.41 [85.09 – 169.98] | -49.0% (-65.6% to -32.4%) | <0.001* |
| Embu | 7 (3.3) | 145.03 (77.44 – 212.62) | -44.9% (-66.5% to -9.5%) | 0.018* | 167.28 [57.36 – 203.00] | -27.3% (-65.6% to -32.4%) | 0.004* |
| Homabay | 4 (1.9) | 137.38 (52.68 – 222.07) | -47.8% (-72.5% to -1.1%) | 0.046† | 153.80 [100.35 – 174.40] | -33.1% (-65.6% to -32.4%) | 0.015† |
| Kakamega | 4 (1.9) | 118.48 (45.44 – 191.53) | -55.0% (-76.3% to -14.7%) | 0.014† | 117.96 [113.55 – 123.42] | -48.7% (-65.6% to -32.4%) | 0.002† |
| Kiambu | 20 (9.4) | 198.78 (143.97 – 253.58) | -24.5% (-45.4% to 4.4%) | 0.089 | 205.67 [127.03 – 265.50] | -10.6% (-39.6% to 18.4%) | 0.073 |
| Kilifi | 47 (22.0) | 174.34 (142.98 – 205.69) | -33.8% (-48.4% to -15.2%) | 0.001* | 171.17 [124.37 – 200.67] | -25.6% (-37.8% to -13.4%) | <0.001* |
| Kirinyaga | 4 (1.9) | 206.33 (79.12 – 333.53) | -21.7% (-58.7% to 48.5%) | 0.455 | 184.77 [80.33 – 332.32] | -19.7% (-37.8% to -13.4%) | 0.47 |
| Kisumu | 14 (6.5) | 209.88 (140.71 – 279.04) | -20.3% (-45.0% to 15.5%) | 0.231 | 112.76 [65.08 – 341.85] | -51.0% (-107.5% to 5.5%) | 0.046* |
| Machakos | 19 (8.9) | 213.26 (152.93 – 273.59) | -19.0% (-41.8% to 12.7%) | 0.211 | 204.87 [131.23 – 269.66] | -10.9% (-38.9% to 17.0%) | 0.131 |
| Migori | 2 (0.9) | 89.12 (11.42 – 166.83) | -66.2% (-86.1% to -17.7%) | 0.017† | 89.13 [88.32 – 89.93] | -61.3% (-38.9% to 17.0%) | 0.013† |
| Muranga | 3 (1.4) | 228.62 (65.87 – 391.38) | -13.2% (-58.3% to 80.5%) | 0.705 | 274.46 [128.09 – 283.32] | 19.3% (-38.9% to 17.0%) | 0.848 |
| Nakuru | 4 (1.9) | 195.03 (74.79 – 315.27) | -26.0% (-60.9% to 40.4%) | 0.357 | 213.84 [87.21 – 302.85] | -7.00% | 0.591 |
| Nyeri | 10 (4.7) | 301.44 (183.90 – 418.98) | 14.5% (-25.2% to 75.2%) | 0.534 | 160.43 [82.56 – 236.94] | -30.3% (-111.5% to 51.0%) | 0.073 |
| Trans Nzoia | 5 (2.3) | 146.06 (65.52 – 226.60) | -44.5% (-68.9% to -1.2%) | 0.045† | 113.79 [111.67 – 140.14] | -50.5% (-111.5% to 51.0%) | 0.038† |

\* Statistically significant at the 5% level ( $p < 0.05$ )
\*\* Confidence interval and p-value not reported due to small sample size ( $n=1$ )
\*\*\* Confidence interval calculated under model assumptions despite a single observation. Interpret with caution.
† Interpret p-value with caution due to small sample size ( $n \leq 5$ )

Children aged five years and above had lower median costs (26%) than the reference (<1 year), while patients from urban settings faced up to 43% higher costs than those from rural areas. Care at Level 4 hospitals was also associated with lower mean and median costs compared with Level 5 facilities.

Referred patients had a higher median cost of treatment than direct admissions. Diagnosis type also had a strong influence on treatment costs, were patients with multiple co-morbidities reporting significantly higher costs. Details on specific comorbidities and their frequency are provided in Figure S1 under Appendix 9.

Geographical variation was also evident. Several counties, including Busia, Kakamega, Kilifi, Homabay, Migori, Kisumu, and Trans Nzoia, recorded significantly lower costs compared with Nairobi (reference). All the differences in costs described in this section were statistically significant.

### Sensitivity analysis

The tornado diagram in Figure 2 presents the results of the sensitivity analysis and highlight’s the parameters that the cost of treatment was most sensitive to. Varying staff time by applying the derived adjustment factor increased median costs to USDLJ305.37 and USDLJ330.89 from the healthcare provider and societal perspectives, respectively. Varying the hospital bed costs had a smaller effect, were the lower limit reduced the median costs by 8-10% and the upper limit increased it by 6-10%. The effect of the parameters obtained from literature were extremely low. The detailed results are presented in Table S10 under Appendix 10.

**Figure 2:**
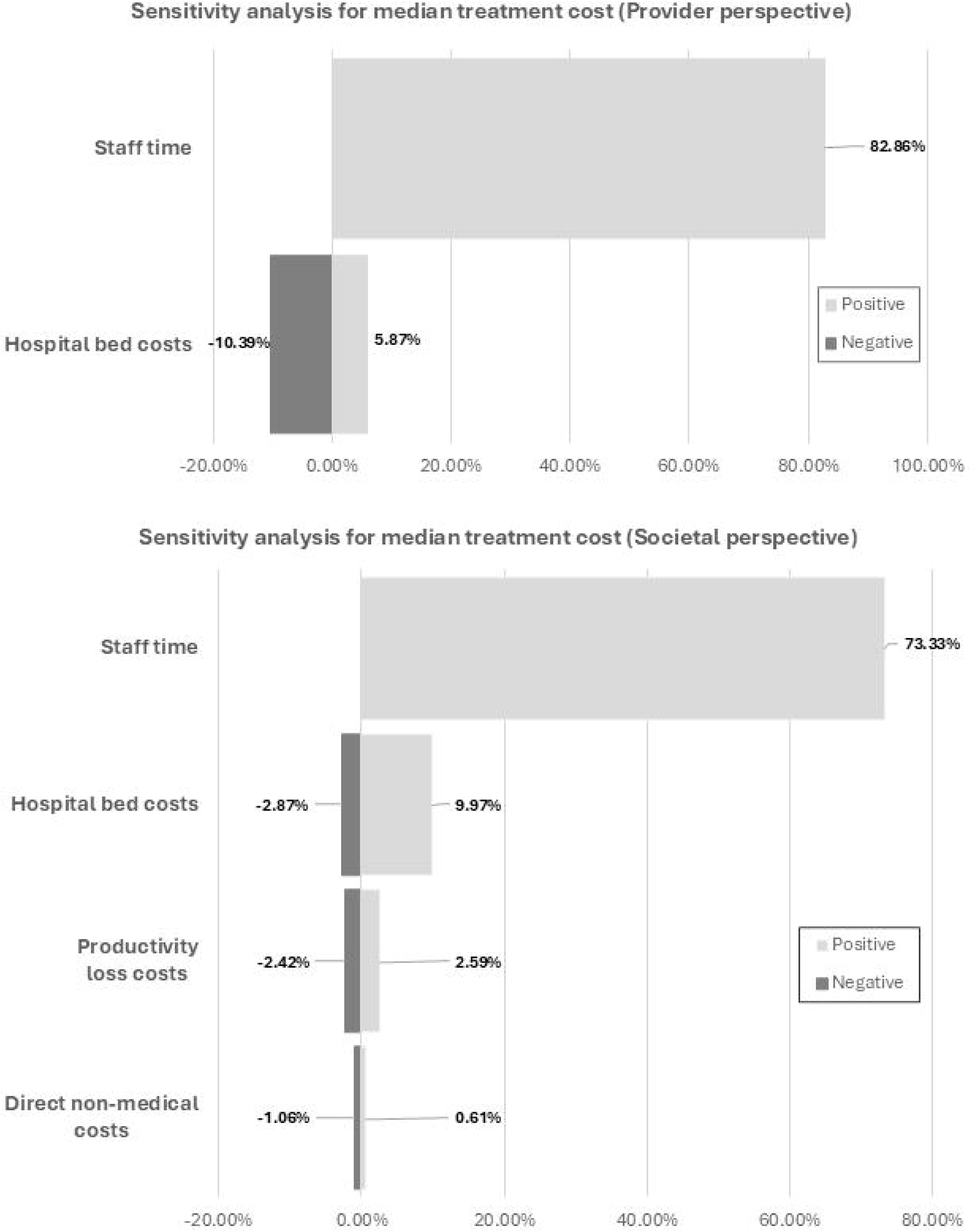
Tornado diagrams showing the results of the one-way sensitivity analyses.

### Catastrophic Health Expenditure

The projected occurrence of CHE among households was present at all levels of OOP expenditure in rural areas. At the national average for OOP health expenditure, at least 2 households from the rural setting and 1 household from urban areas were projected to face CHE. As the share of treatment costs paid OOP reached 60%, the occurrence of CHE significantly increased with households from all wealth quintiles potentially being affected (rural setting). If patients had to pay for the full treatment OOP, then up to 53% of rural households and close to 70% of urban households could face CHE. This is detailed in Table 5.

**Table 5:** Catastrophic health expenditure.

|  | Quintile | Total number of patients | OOP share (20%) | OOP share (24.2%) | OOP share (40%) | OOP share (60%) | OOP share (80%) | OOP share (100%) |
| --- | --- | --- | --- | --- | --- | --- | --- | --- |
| **Rural** | Poorest (Q1) | 138 | 1 (.7%) | 2 (1.4%) | 8 (5.8%) | 23 (16.7%) | 50 (36.2%) | 73 (52.9%) |
|  | Poorer (Q2) |  | 1 (.7%) | 1 (.7%) | 3 (2.2%) | 14 (10.1%) | 23 (16.7%) | 41 (29.7%) |
|  | Middle (Q3) |  | 1 (.7%) | 1 (.7%) | 2 (1.4%) | 3 (2.2%) | 14 (10.1%) | 23 (16.7%) |
|  | Richer (Q4) |  | 0 (0%) | 1 (.7%) | 1 (.7%) | 2 (1.4%) | 3 (2.2%) | 10 (7.2%) |
|  | Richest (Q5) |  | 0 (0%) | 0 (0%) | 0 (0%) | 1 (.7%) | 1 (.7%) | 1 (.7%) |
| **Urban** | Poorest (Q1) | 76 | 1 (1.3%) | 1 (1.3%) | 2 (2.6%) | 15 (19.7%) | 35 (46.1%) | 52 (68.4%) |
|  | Poorer (Q2) |  | 0 (0%) | 0 (0%) | 0 (0%) | 1 (1.3%) | 1 (1.3%) | 2 (2.6%) |
|  | Middle (Q3) |  | 0 (0%) | 0 (0%) | 0 (0%) | 0 (0%) | 1 (1.3%) | 1 (1.3%) |
|  | Richer (Q4) |  | 0 (0%) | 0 (0%) | 0 (0%) | 0 (0%) | 0 (0%) | 0 (0%) |
|  | Richest (Q5) |  | 0 (0%) | 0 (0%) | 0 (0%) | 0 (0%) | 0 (0%) | 0 (0%) |

## Discussion

The purpose of the study was to provide the first estimates for treating a hospitalised measles paediatric patient in Kenya and assess the financial strain the disease places on households, addressing key evidence gaps relevant to measles control and the financial burden the disease places on a patient’s family and healthcare system. Patient level data from 16 public hospitals was extracted and analysed. By adopting a healthcare provider and societal perspective, the findings provided a comprehensive estimate of the treatment costs. The median cost of treatment from the healthcare provider perspective was USD 139.11 (mean: USD 155.06), while the median estimate from the societal perspective was 27% higher (median: USD176.18; mean: USD 205.11).

A comparison of measles treatment costs from other LMICs found four studies that focused on hospitalised children [12,53–55]. The Ugandan study reported a similar cost of treatment (7% higher) from the provider perspective of USD 150 (in 2025 USD), and a study conducted in Bangladesh reported a slightly lower (20%) societal cost (adjusted to 2025 equivalents) of USD 146 [53,54]. However, two other studies conducted in LMICs reported much lower costs [12,55], which could be attributed to the costing methodology, perspective and context specific prices of items. Measles infection can lead to suppression of the immune system, leading to secondary infections [56]. Variation in treatment protocols for managing secondary infections across countries, can further contribute to the cost differences.

The two main drivers of the cost of treatment were the staff-related costs and hospital bed costs. Staff time was the highest cost driver, given the multi-cadre approach required for treating measles and the labour-intensive nature of the disease. In Kenya, the average income for healthcare workers is three folds higher than that of the general population [49,57]. A study estimating the treatment cost of measles in Uganda also reported human resource related costs to be the main cost driver [53]. The relatively higher costs associated with bed charges includes the cost of accommodation, meals and nursing services [58]. A more severe form of the disease or delays in decision making could lead to an increased duration of hospitalization, which in turn increases the total hospital bed costs. A study by Onyango et al. attributed hospital related treatment delays in Kenyan public hospitals to severity of disease on presentation, slow triage, inadequate staffing and long administrative processes [59]. Task shifting across cadres and improving patient flow could enhance efficiency, leading to shortening of hospital stay without compromising on the quality of care provided.

The results of the sub-group analyses highlighted how costs significantly differ among the different groups. Children under one year faced the highest treatment costs, with the 7 of 10 cost categories in this age-group being the highest when compared to the other age-groups. Patients from urban settings experienced greater costs, that could be explained by the higher direct non-medical and indirect costs when compared to their rural counterparts. The higher costs experienced by referred cases, could be associated to more severe cases as patients in these sub-groups had a longer hospitalisation period (Median: 7 days), and reported higher costs for medications and laboratory tests when compared to direct admissions. Patients that received care at lower level hospitals (level 4) had significantly lower costs, which could be explained with the more specialised and intensive care that is provided at higher level facilities [60]. Comorbidities significantly affected the cost, with higher costs reported as the number of comorbidities increased. The most common comorbidities were pneumonia (n=110), gastrointestinal disorders (n=73) and dehydration (n=56). In terms of counties, patient from Nairobi reported the highest cost when compared to other counties, likely due to the higher non-medical and indirect costs, greater intensity of care and longer duration of hospitalization (7 days).

The sensitivity analysis highlighted how our assumptions around health care workers affected the total cost. When staff time was adjusted to account for staff shortages, the cost increased drastically (73-83%). Staffing gaps in public hospitals across Africa are common, with patients often receiving reduced quality of care as healthcare workers are overworked [61,62]. As a result, the amount of time spent with a patient could be less than what is required, leading to an underestimation of costs. The effect of hospital bed costs was less pronounced, as bed charges in public hospitals in Kenya are generally similar, which is reflected in the narrow IQR.

The financial burden on households and the risk of facing CHE were substantial. Using simulated household expenditure scenarios, rural households from most wealth quintiles (Q3–Q5) could face catastrophic health spending even when their out-of-pocket share was below the national average of 24.2%. More than half of households, irrespective of geographical setting, were at risk of falling into poverty if required to pay the total cost out of pocket. These findings suggest that policymakers would need to cover a majority of treatment costs (>80%) to shield households from impoverishment. This could be achieved by through public health financing mechanisms, such as improving insurance coverage, targeted subsidies, dedicated disease-specific support for high-burden areas and improving vaccine coverage to help reduce disease transmission.

One key limitation of this study was that pre and post hospitalization costs were not captured. These costs are important as patients with measles could suffer from long-term complications that require post discharge care [63]. Patient were assumed to have been diagnosed with measles based on clinical assessment rather than confirmatory tests as no variable in the CIN dataset described any standard laboratory tests used to confirm a measles diagnosis. Another limitation was that we did not collect household level cost data for the CHE analysis and used a simulation model that relied on hypothetical scenarios. This may not have captured the real-world variations in household spending and coping strategies.

The studies had many strengths, as it provided the first estimate for the cost of treating hospitalised children with measles in Kenya. Patient level data from 16 hospitals of different levels were included in the study, which gave a good representation of both service levels and geographic variation in costs and treatment modalities seen in urban and rural hospitals. The use of local prices and primary data for patients provided a context specific estimate. Including both the healthcare provider and societal perspective, provided a broader understanding on the financial burden of the disease. The inclusion of a CHE analysis further highlighted the financial distress households could face due to the disease.

Future studies could build on this work by collecting patient level data on pre and post hospitalization care, especially for those patients that suffer for long-term complications due to the disease. Time and motion analyses should be conducted to determine the amount of time different cadres spend in measles care, as this would provide a more accurate estimate of staff times. Surveys of household of patients treated for measles should be conducted to get a better understanding on the incidence of CHE across different regions and income groups. Expanding the study population to include patients of all ages, different care settings and those treated at private facilities would provide a more complete picture on the economic burden of the disease.

## Conclusion

The economic burden of measles among hospitalised paediatric patients in Kenya is substantial, with staff and hospital bed costs being the primary drivers. Younger children, patients from urban settings and those referred reported significantly higher costs, and the same trend was seen in patients from higher level hospitals, those with multiple comorbidities and admitted in Nairobi. Another key finding was that a large proportion (>50%) of households could face CHE if the full payment for treatment was made OOP. Researchers can use these outputs to inform economic evaluations of difference vaccination strategies for measles vaccine in Kenya or similar settings. The findings can also be used by hospitals to improve efficiency around the main costs drivers to help reduce treatment expenses. At the sub-national level, policy makers can use the evidence to plan health budgets, while at the national level, it provides a justification for increased investment to improve both insurance and vaccine coverage levels to help reduce the health and economic burden of measles in Kenya.

## Data Availability

All data produced in the present study are available upon reasonable request to the authors

## Data availability statement

The data will be shared on reasonable request to the corresponding author.

## Contributions

EB, PM, SO, FM, and OO conceptualised the study. PM led the data analysis and was supported by SO, IW, and FO. PM drafted the initial manuscript which was subsequently revised for important intellectual content by all authors. All authors read and approved the final manuscript. The work was supervised by FO, FM, OO, SO, and EB. While EB, SO, FM, and OO obtained the funding for this work. PM is responsible for the overall content as guarantor.

## Ethics approval

Ethical approval was obtained from the KEMRI Scientific and Ethical Review Committee, reference number KEMRI/SERU/CGMR-C/320/4983. All experts provided verbal consent before being interviewed, and participation was voluntary. Responses were anonymised to protect confidentiality.

