## Supplementary_Materials for "Economic Burden of Measles among Hospitalised Children in Kenya"

### Appendix 1: Table containing hospital bed costs for public hospitals in Kenya.

Table S1: Unit costs for hospital bed charges in Kenyan public hospitals

| **Select Hospital** | **Hospital level** | **Daily Bed Charges**  **(KES)** | **Daily Bed Charges**  **(USD)** |
| --- | --- | --- | --- |
| Mbagathi County Hospital | Level 5 | 1000 | 7.74 |
| Naivasha Level 5 Hospital | Level 5 | 650 | 5.03 |
| Busia County Referral Hospital | Level 5 | 200 | 1.55 |
| Kisumu County Hospital | Level 5 | 750 | 5.80 |
| Nakuru Level 5 Hospital | Level 5 | 500 | 3.87 |
| Jaramogi Oginga Odinga Teaching and Referral Hospital | Level 5 | 1000 | 7.74 |
| Kiambu Level 5 Hospital | Level 5 | 800 | 6.19 |
| Machakos Level 5 Hospital | Level 5 | 200 | 1.55 |
| Kakamega County General Teaching and Referral Hospital | Level 5 | 600 | 4.64 |
| Nyeri County Referral Hospital | Level 5 | 1900 | 14.70 |
| Embu Level 5 Teaching and Referral Hospital | Level 5 | 100 | 0.77 |
| Bungoma County Referral Hospital | Level 5 | 550 | 4.25 |
| Thika Level 5 Hospital | Level 5 | 250 | 1.93 |
| Machakos Level 5 Hospital | Level 5 | 800 | 6.19 |

### Appendix 2: Table of unit costs for laboratory and radiology

Table S2: Unit costs for laboratory and radiology tests

| **Test name** | **Hospital 1** | **Hospital 2** | **Hospital 3** | **Hospital 4** | **Hospital 5** | **Hospital 6** | **Hospital 7** | **Others** | **Median Unit Cost**  **(KES, 2025)** | **Median Unit Cost**  **(USD, 2025)** | **Source** |
| --- | --- | --- | --- | --- | --- | --- | --- | --- | --- | --- | --- |
| **Malaria test** |  |  |  |  |  |  |  |  |  |  |  |
| Blood slide | 100.00 | 100.00 | 50.00 | 100.00 | 100.00 | 50.00 | 100.00 |  | 110.82 | 0.86 | Public health facilities in 2023 |
| Both (B/slide + Rapid) | 200.00 | 200.00 | 100.00 | 200.00 | 200.00 | 100.00 | 200.00 |  | 221.64 | 1.71 | Public health facilities in 2023 |
| **Haematology test** |  |  |  |  |  |  |  |  |  | ` |  |
| Full Hemogram | 600.00 | 1000.00 | 500.00 | 500.00 | 400.00 | 500.00 | 500.00 |  | 554.10 | 4.29 | Public health facilities in 2023 |
| Hb | 200.00 | 200.00 | 100.00 | 100.00 | 200.00 | 150.00 | 150.00 |  | 174.15 | 1.35 | Public health facilities in 2023 |
| **RBS** |  |  |  |  |  |  |  |  |  |  |  |
| Strip | 150.00 | 200.00 | 150.00 | 100.00 | 100.00 | 150.00 | 150.00 |  | 166.23 | 1.29 | Public health facilities in 2023 |
| Laboratory |  |  |  |  |  | 400.00 |  |  | 443.28 | 3.43 | Public health facilities in 2023 |
| **Chemistry** |  |  |  |  |  |  |  |  |  |  |  |
| Sodium & Potassium | 600.00 | 1000.00 | 400.00 | 1000.00 |  | 800.00 |  |  | 886.56 | 6.86 | Public health facilities in 2023 |
| Urea & Creatinine | 600.00 | - | 400.00 | 800.00 |  | 800.00 | 1000.00 |  | 886.56 | 6.86 | Public health facilities in 2023 |
| Calcium | 500.00 | 400.00 | 500.00 | 200.00 | 200.00 | 500.00 | 500.00 |  | 554.10 | 4.29 | Public health facilities in 2023 |
| Albumin | 200.00 | 300.00 | 200.00 |  |  | 400.00 |  |  | 318.61 | 2.46 | Public health facilities in 2023 |
| LFT | 1600.00 | 1000.00 | 1600.00 | 2000.00 | 1000.00 | 3000.00 | 1500.00 |  | 1773.13 | 13.72 | Public health facilities in 2023 |
| U/E/C | 1500.00 | 1600.00 | 1000.00 | 1000.00 | 200.00 | 2000.00 | 1500.00 |  | 1662.31 | 12.86 | Public health facilities in 2023 |
| **HIV** |  |  |  |  |  |  |  |  |  |  |  |
| Strip |  |  |  | 100.00 | 400.00 |  |  |  | 360.17 | 2.79 | Public health facilities in 2023 |
| PCR |  |  | 1000.00 |  |  |  |  |  | 1108.20 | 8.57 | Public health facilities in 2023 |
| **Microbiology** |  |  |  |  |  |  |  |  |  |  |  |
| Lumbar puncture | 1000.00 | 1000.00 | 500.00 | 1200.00 | 600.00 | 1000.00 |  |  | 1108.20 | 8.57 | Public health facilities in 2023 |
| Blood Culture | 2000.00 | 1500.00 | 2500.00 | 2000.00 | 1000.00 | 1000.00 | 1000.00 |  | 1741.46 | 13.47 | Public health facilities in 2023 |
| **X-Ray** |  |  |  |  |  |  |  |  |  |  |  |
| Chest | 600.00 | 600.00 | 1040.00 | 400.00 | 550.00 |  | 400.00 |  | 443.22 | 3.43 | Public health facilities in 2023 |
| Wrist | 600.00 | 600.00 |  |  |  |  |  |  | 664.92 | 5.14 |  |
| Others | 800.00 | 600.00 | 1040.00 |  |  |  |  |  | 901.34 | 6.97 | Public health facilities in 2023 |
| **Urine** |  |  |  |  |  |  |  |  |  |  |  |
| Urinalysis | 200.00 | 300.00 | 100.00 | 150.00 | 150.00 | 150.00 | 200.00 |  | 197.89 | 1.53 | Public health facilities in 2023 |
| **TB Test** |  |  |  |  |  |  |  |  |  |  |  |
| Mantoux |  |  |  |  |  |  |  | 1115.2 | 1115.25 | 8.63 | https://cerbalancetafrica.ke/media/yj4k0so1/plk-price-catalogue-2022.pdf |

*Hospital 1: Kakamega County Referral Hospital, Hospital 2: Machakos Level 5 Hospital, Hospital 3: Bungoma County Referral Hospital, Hospital 4: JOOTRH, Hospital 5: Mama Lucy K Hospital, Hospital 6: Kitale County Hospital, Hospital 7: Kisumu County Hospital, and Others: Cerba Lancet Kenya.*

### Appendix 3: Table containing the cost of blood transfusion from the survey

Table S3: Survey-derived unit cost for blood transfusion

|  | **Hospital 1 (KES)** | **Hospital 2 (KES)** | **Hospital 3 (KES)** |
| --- | --- | --- | --- |
| **Cost of laboratory tests** | 500 | 500 | 500 |
| **Cost of consumables** | 200 | 500 | 500 |
| **Total cost of blood transfusion** | 700 | 1000 | 1000 |
| **Median cost (KES)** | 1000 |  |  |
| **Median cost (USD)** | 7.74 |  |  |
| **Key** |  |  |  |
| Hospital 1 = Port Reitz Sub County Hospital | |  |  |
| Hospital 2 = Jaramogi Oginga Odinga Teaching and Referral Hospital | | | |
| Hospital 3 = Nyanyuki Teaching and Referral Hospital | |  |  |

### Appendix 4: Table containing staff times from survey.

Table S4: Expert opinion-based estimates of staff time per cadre

| **Cadre** | **Unit** | **Staff 1** | **Staff 2** | **Staff 3** | **Staff 4** | **Mean** |
| --- | --- | --- | --- | --- | --- | --- |
| Specialist time (first visit) | Minutes | 30 | 15 | 20 | 20 | 21 |
| Specialist time (subsequent visit) | Minutes | 20 | 15 | 20 | 10 | 16 |
| Medical officer (First visit) | Minutes | 10 | 10 | 10 | - | 10 |
| Medical officer (subsequent visit) | Minutes | 5 | 5 | 5 | - | 5 |
| Clinical officer (first visit) | Minutes | 15 | 10 | 10 | - | 11 |
| Clinical officer (subsequent visit) | Minutes | 15 | 5 | 10 | - | 9 |
| Nurse time per day with patient | Minutes | 60 | 64 | 30 | - | 49 |
| Nutritionist | Minutes | 20 | 20 | - | - | 20 |

### Appendix 5: Justification for selection of reference categories in subgroup

| **Subgroup** | **Reference Category** | **Justification Summary** |
| --- | --- | --- |
| Gender | Female | Common practice in health studies |
| Age-group | Under 1 year | Focus on vulnerable younger children |
| Geographical setting | Rural | Standard in health economic studies done in LMIC |
| Hospital Level | Level 5 | The largest group |
| Referral status | Not referred | The larger group |
| Diagnosis | Measles only | Simplest cases for comparison |
| County | Nairobi | County with the largest sample size for stability |

Table S5: Justification for selection of reference categories

### Appendix 6: Adjusted staff time estimates

Table S6: Adjusted staff time estimates

| **Cadre of Staff** | **Base Case (minutes)** | **Average Number of healthcare workers at a level 5 hospital [KHFA]**  **(Number of staff)** | **Norms and Standards (Number of staff)** | **Adjustment Factor (Norms and Standards/Average KHFA)** | **Adjusted Value (minutes)** |
| --- | --- | --- | --- | --- | --- |
| Pediatrician (first visit) | 21 | 2.3 | 9.0 | 3.91 | 81 |
| Pediatrician (subsequent visit) | 16 | 2.3 | 9.0 | 3.91 | 61 |
| Medical officer (First visit) | 10 | 23.1 | 45.0 | 1.95 | 19 |
| Medical officer (subsequent visit) | 5 | 23.1 | 45.0 | 1.95 | 10 |
| Clinical officer (first visit) | 11 | 35.3 | 30.0 | 0.85 | 10 |
| Clinical officer (subsequent visit) | 9 | 35.3 | 30.0 | 0.85 | 8 |
| Nurse | 49 | 19.8 | 20.0 | 1.01 | 49 |
| Nutritionist | 20 | 5.9 | 5.0 | 0.85 | 17 |

### Appendix 7: Detailed methodology for catastrophic healthcare expenditure analysis

The methodology for the CHE analysis was directly adapted from that proposed by Mandaliya et al. and is detailed in this section [1].

The incidence of catastrophic health expenditure (CHE) for each patient was estimated by using a simulation-based approach. Thirty unique scenarios for urban and rural settings were generated by varying the possible amount of the total cost a patient’s household could have paid out of pocket (OOP) and the wealth quintile the household would belong to, and individual patients were then simulated across each scenario. If a patient’s household was determined to have incurred CHE, then it was scored one, otherwise zero. The number of households that could have incurred CHE were summed across each scenario.

Standard CHE analysis utilises patient level data for both OOP expenditure and non-food household expenditure [2]. As patient level data for these parameters were unavailable, the values were derived using data obtained from national surveys and published literature. Specifically, for non-food household expenditure (NFHE), the secondary sources were used to calculate the value, while for OOP expenditure, the proportion of treatment costs that could have been paid OOP were varied between 20-100%. All monetary values obtained from literature were first converted to their 2025 KES equivalents.

**1. Calculating projected out-of-pocket expenditure**

To estimate the total OOP amount paid by each patient (p) we first obtained the total cost for each patient:

$${TotalCost}_{p} = {DirectMedicalCost}_{p} + {DirectNonMedicalCost}_{p}$$

For each OOP share scenario, the projected expenditure was calculated as:

$${OOP}_{p,s}= {TotalCost}_{p} \times Shares$$

The specific *Share_s_* values were 0.20, 0.242, 0.40, 0.60, 0.80, and 1.00. The value of 0.242 was chosen as a baseline, as this is the currently reported average OOP health expenditure for Kenya [3].

**2. Estimation of annual non-food household expenditure**

The denominator, which is the annual NFHE, was determined for each wealth quintile (q) as per the steps described below:

**Step 1: Mean adult equivalent (AE) expenditure**

The mean adult equivalent (AE) expenditure and its percentage shared for each quintile for both urban and rural areas were sourced from the Kenya Poverty Report 2022 [4]. Mean AE expenditure was distributed across quintiles (Q1–Q5) by multiplying the mean total AE with the reported quintile percentage shares. The value for each quintile (q) was calculated as:

$${AE}_{q} = {AE}_{mean} \times\left( \frac{{Proportion}_{q}}{20} \right)$$

where *AE_q_* is the mean AE expenditure for quintile *q*, and *Proportion_q_* is the quintile’s proportion of total expenditure.

**Step 2: Non-food adult equivalent expenditure**

Non-food AE expenditure (NFAE) for each quintile was calculated by multiplying the resulting *AE_q_* by the national average non-food expenditure percentage, obtained from the Kenya Poverty Report 2022 [4]:

$${NFAE}_{q} = {AE}_{q} \times Non-food expenditure (\%)$$

**Step 3: Weighted AE factor**

To adjust for household composition, we applied adult equivalent weights to the national age-group proportions (0–4, 5–14, and 15+ years) derived from the 2019 Kenya National Population and Housing Census [5], and AE weights for each age group from the Kenya Poverty Report 2022 [4]. A weighted AE factor (WAF) was calculated as:

$$WAF = ({Weight}_{0-4} \times{Proportion}_{0-4}) + ({Weight}_{5-14} \times{Proportion}_{5-14}) + ({Weight}_{15+} \times P{ropotion}_{15+})$$

**Step 4: AE per household**

The adult equivalent household size (AE_HH_) was calculated as:

$${AE}_{HH} = Household Size x WAF$$

The average household size was obtained from the Kenya demographic and health survey 2022 [6].

**Step 5: Monthly and Annual non-food household expenditure**

The monthly NFHE was calculated by geographical setting across the different wealth quintiles:

$${NFHE}_{monthly,q} = {AE}_{HH} \times{NFAE}_{q}$$

Which was converted to the annual NFHE:

$${NFHE}_{annual, q} = {NFHE}_{monthly,q} \times12$$

**3. Determination of catastrophic health expenditure (CHE)**

We then calculated the CHE ratio (*R_p,s_*) for each patient (*p*) across the different unique scenarios as follows:

$$R_{p,s}=\frac{{OOP}_{p,s}}{{NFHE}_{annual, q}}$$

A household was classified to have incurred CHE if:

$$R_{p,s} > 0.4$$

The total number households experiencing CHE under each unique scenario were obtained the number of individual households experiencing CHE per scenario.

Table S7 lists the data sources for the parameters and Table S8 summarizes the derived NFHE by quintile and geographical setting.

Table S7: Parameter inputs used to derive non-food household expenditure

| **Parameter name** | **Rural** | **Urban** | **Notes** | **Source** |
| --- | --- | --- | --- | --- |
| **Mean AE expenditure (KES, 2025)** | 6,717 | 15,261 | Converted reported value to 2025, KES. | Kenya Poverty Report 2022 [4] |
| **Quintile shares of consumption (used to derive quintile means)**  *Poorest (Q1) to Richest (Q5)* | 8.5%, 11.4%, 14.4%, 20.4%, 45.3% | 3.7%, 11.5%, 15.9%, 25.5%, 43.4% |  | Kenya Poverty Report 2022 [4] |
| **Non-food %** | 34.2 | 55.2 |  | Kenya Poverty Report 2022 [4] |
| **Household (HH) size**  **(persons)** | 4.41 | 3.12 |  | Kenya demographic and health survey 2022 [6] |
| **National Age proportions by age group  (0–4 / 5–14 / 15+)** | 12.7 / 28.9 / 58.4 | 12.3 / 20.8 / 66.9 |  | 2019 Kenya population and housing census [5] |
| **AE weights by age-group  (0–4 / 5–14 / 15+)** | 0.24 / 0.65 / 1.0 | 0.24 / 0.65 / 1.0 |  | Kenya Poverty Report 2022 [4] |
| **Income per month  per earner (15+) (2025, KES)** | Low: 9,052 Average: 21,053 High: 33,783 | Low: 16,967 Average: 26,641 High: 38,287 | Converted reported value to 2025, KES. | Economic Survey 2024 [7] |
| **Employment to population ratio, 15+, total (%) (modeled ILO estimate) - Kenya** | 63 | 63 | 2024 national value | World Bank Open Data [8] |

Table S8: Derived non-food household expenditure by quintile and residence type (monthly and annual)

| **Residence type** | **Quintile** | **Mean AE expenditure (KES, 2025)** | **Proportion (%)** | **Total expenditure  per month per AE (KES, 2025)** | **Non-food proportion (%)** | **Non-food per AE  per month (KES, 2025)** | **HH  size  (persons)** | **Proportion per  age-group  (0–4 / 5–14 / 15+)** | **AE weights by  age-group  (0–4 / 5–14 / 15+)** | **Weighted AE**  **factor** | **AE  per  HH** | **Non-food expenditure per household per month  (KES, 2025)** | **Non-food expenditure per household per year  (KES, 2025)** |
| --- | --- | --- | --- | --- | --- | --- | --- | --- | --- | --- | --- | --- | --- |
| **Rural** | Q1 | 6717 | 8.5 | 2,855 | 34.2 | 976 | 4.41 | 12.6 / 26.3 / 61.1 | 0.24 / 0.65 / 1.0 | 0.80233 | 3.54 | 3,454 | 41,454 |
|  | Q2 |  | 11.4 | 3,829 |  | 1309 |  |  |  |  |  | 4,633 | 55,597 |
|  | Q3 |  | 14.4 | 4,836 |  | 1654 |  |  |  |  |  | 5,852 | 70,227 |
|  | Q4 |  | 20.4 | 6,851 |  | 2343 |  |  |  |  |  | 8,291 | 99,489 |
|  | Q5 |  | 45.3 | 15,214 |  | 5203 |  |  |  |  |  | 18,410 | 220,924 |
| **Urban** | Q1 | 15261 | 3.7 | 2,823 | 55.2 | 1558 | 3.12 | 12.6 / 26.3 / 61.1 | 0.24 / 0.65 / 1.0 | 0.83372 | 2.60 | 4,054 | 48,646 |
|  | Q2 |  | 11.5 | 8,775 |  | 4844 |  |  |  |  |  | 12,600 | 151,198 |
|  | Q3 |  | 15.9 | 12,132 |  | 6697 |  |  |  |  |  | 17,421 | 209,048 |
|  | Q4 |  | 25.5 | 19,458 |  | 10741 |  |  |  |  |  | 27,939 | 335,265 |
|  | Q5 |  | 43.4 | 33,116 |  | 18280 |  |  |  |  |  | 47,551 | 570,608 |

### Appendix 8: Sub-group analysis (provider perspective)

Table S9: Sub-group analysis from the healthcare provider perspective

| **Name** | **N (%)** | **Mean total cost (95% CI)** | | | **Median total cost [IQR]** | | |
| --- | --- | --- | --- | --- | --- | --- | --- |
|  |  | **2025, USD** | **% Difference (CI)** | **P-value** | **2025, USD** | **% Difference (CI)** | **P-value** |
| **Gender** | | | | | | | |
| Female | 79 (36.9) | 163.81  (133.59 to 194.02) | Reference | - | 148.08  [93.74, 191.25] | Reference | - |
| Male | 135 (63.1) | 149.94  (128.78 to 171.09) | -8.47%  (-29.72% to 12.79%) | 0.435 | 132.33  [90.66, 184.16] | -10.64%  (-24.70% to 3.42%) | 0.146 |
| **Age-Group** | | | | | | | |
| Under 1 year | 74 (34.6) | 156.74  (128.17 to 185.3) | Reference | - | 148.29  [104.97, 200.01] | Reference | - |
| 1-4 years | 110 (51.4) | 163.05  (138.68 to 187.43) | 4.03%  (-20.49% to 28.55%) | 0.748 | 139.11  [90.01, 184.16] | -6.19%  (-21.17% to 8.78%) | 0.181 |
| 5 years and older | 30 (14.0) | 121.60  (86.79 to 156.41) | -22.42%  (-48.75% to 3.91%) | 0.095 | 111.82  [73.02, 154.09] | -24.60%  (-48.57% to -0.62%) | 0.021* |
| **Geographical setting** | | | | | | | |
| Rural | 138 (64.5) | 147.72  (126.95 to 168.48) | Reference | - | 126.78  [87.41, 169.59] | Reference | - |
| Urban | 76 (35.5) | 168.39  (136.49 to 200.28) | 13.99%  (-12.90% to 40.88%) | 0.308 | 152.82  [110.64, 196.06] | 20.54%  (-2.55% to 43.63%) | 0.004* |
| **Hospital level** | | | | | | | |
| Level 5 | 143 (66.8) | 160.47  (139.48 to 181.46) | Reference | - | 142.98  [92.5, 189.35] | Reference | - |
| Level 4 | 70 (32.7) | 138.57  (112.66 to 164.47) | -13.65%  (-33.35% to 6.05%) | 0.175 | 127.06  [90.86, 166.27] | -11.13%  (-27.30% to 5.03%) | 0.195 |
| Level 6 | 1 (0.5) | 535.32  (0 to 1372.68) | 233.60%  (-196.33% to 663.52%) | 0.287 | 535.32  [535.32, 535.32] | 274.40% | -** |
| **Referral status** | | | | | | | |
| Not referred | 191 (89.3) | 150.33  (132.38 to 168.27) | Reference | - | 133.39  [90.01, 183.37] | Reference | - |
| Referred | 23 (10.8) | 194.33  (127.48 to 261.17) | 29.27%  (-17.80% to 76.34%) | 0.223 | 181.74  [132.33, 274.84] | 36.25%  (-9.43% to 81.93%) | 0.003* |
| **Diagnosis** | | | | | | | |
| Measles Only | 27 (12.6) | 95.54  (67.96 to 123.12) | Reference | - | 94.67  [58.47, 126.74] | Reference | - |
| Measles +1 | 45 (21.0) | 143.49  (111.4 to 175.58) | 50.19%  (-4.66% to 105.03%) | 0.073 | 136.31  [90.66, 177.67] | 43.98%  (-5.34% to 93.31%) | 0.010* |
| Measles +2 or more | 142 (66.4) | 170.04  (148.63 to 191.45) | 77.98%  (21.93% to 134.03%) | 0.006* | 148.73  [103.13, 191.08] | 57.10%  (18.88% to 95.32%) | <0.001* |
| **County** | | | | | | | |
| Nairobi | 52 (24.3) | 184.06  (150.88 to 217.25) | Reference | - | 153.68  [125.08, 199.04] | Reference | - |
| Bungoma | 1 (0.5) | 156.81  (0 to 360.7) | -14.80%  (-113.99% to 84.38%) | 0.77 | 156.81  [156.81, 156.81] | 2.04% | -** |
| Busia | 18 (8.4) | 114.55  (79.44 to 149.65) | -37.76%  (-59.89% to -15.64%) | <0.001* | 94.48  [67.89, 135.58] | -38.52%  (-59.62% to -17.43%) | <0.001* |
| Embu | 7 (3.3) | 118.00  (60.01 to 175.99) | -35.89%  (-69.45% to -2.33%) | 0.036* | 150.08  [45.9, 162.86] | -2.34%  (-51.21% to 46.53%) | 0.079 |
| Homabay | 4 (1.9) | 111.58  (39.04 to 184.11) | -39.38%  (-80.27% to 1.52%) | 0.059 | 122.26  [80.28, 142.87] | -20.44%  (-56.60% to 15.71%) | 0.08 |
| Kakamega | 4 (1.9) | 96.98  (33.93 to 160.03) | -47.31%  (-82.86% to -11.76%) | 0.009^ϯ^ | 97.88  [93.48, 100.48] | -36.31%  (-46.57% to -26.04%) | 0.009^ϯ^ |
| Kiambu | 20 (9.4) | 134.29  (95.25 to 173.34) | -27.04%  (-52.00% to -2.08%) | 0.034* | 143.27  [85.43, 177.1] | -6.77%  (-36.47% to 22.93%) | 0.047* |
| Kilifi | 47 (22.0) | 142.01  (115.08 to 168.94) | -22.85%  (-43.03% to -2.66%) | 0.027* | 138.91  [101.44, 166.27] | -9.61%  (-25.92% to 6.69%) | 0.010* |
| Kirinyaga | 4 (1.9) | 166.19  (58.15 to 274.24) | -9.71%  (-70.63% to 51.21%) | 0.755 | 150.36 [65.99, 266.39] | -2.16%  (-105.85% to 101.54%) | 0.799 |
| Kisumu | 14 (6.5) | 163.73  (106.84 to 220.63) | -11.05%  (-45.87% to 23.78%) | 0.534 | 83  [48.08, 276.49] | -45.99%  (-108.59% to 16.60%) | 0.071 |
| Machakos | 19 (8.9) | 158.17  (110.99 to 205.35) | -14.07%  (-44.02% to 15.89%) | 0.357 | 148.94  [99.27, 197.74] | -3.08%  (-34.30% to 28.14%) | 0.413 |
| Migori | 2 (0.9) | 71.92  (5.8 to 138.03) | -60.93%  (-97.53% to -24.32%) | 0.001^ϯ^ | 71.92  [71.11, 72.72] | -53.20%  (-60.33% to -46.08%) | 0.031^ϯ^ |
| Murang'a | 3 (1.4) | 190.40  (47.47 to 333.33) | 3.44%  (-76.42% to 83.31%) | 0.933 | 228.6  [105.15, 237.45] | 48.75%  (-14.38% to 111.88%) | 0.604 |
| Nakuru | 4 (1.9) | 135.02  (47.24 to 222.8) | -26.64%  (-76.13% to 22.85%) | 0.291 | 148.38  [59.94, 210.11] | -3.45%  (-81.97% to 75.07%) | 0.611 |
| Nyeri | 10 (4.7) | 257.30  (151.51 to 363.09) | 39.79%  (-22.97% to 102.55%) | 0.214 | 126.02  [65.36, 191.08] | -18.00%  (-91.96% to 55.97%) | 0.168 |
| Trans Nzoia | 5 (2.3) | 116.25  (48.65 to 183.85) | -36.84%  (-75.29% to 1.61%) | 0.06 | 90.86  [88.73, 111.48] | -40.88%  (-98.79% to 17.04%) | 0.059 |

** Statistically significant at the 5% level (p < 0.05)*

*** Confidence interval and p-value not reported due to small sample size (n = 1)*

****Confidence interval calculated under model assumptions despite a single observation. Interpret with caution.*

*† Interpret p-value with caution due to small sample size (n ≤ 5)*

***NB:*** *For categories where the lower bound of the 95% confidence interval (CI) for cost was negative, the lower limit was set to 0, as negative costs are not realistic.*

### Appendix 9: Distribution of comorbidities among hospitalized paediatric patients with measles

|  |  |
| --- | --- |
| Viral Infections | 21 |
| Bacterial Infections | 126 |
| Parasitic Infections | 28 |
| Nutritional and Hematologic Disorders | 36 |
| Respiratory and Allergic Disorders | 9 |
| Gastrointestinal and Hydration Disorders | 129 |
| Neurological and Developmental Disorders | 18 |
| Dermatologic Conditions | 19 |
| Other Conditions | 19 |

Figure S1: Percentage distribution of different comorbidities in hospitalized paediatric patients with measles

### Appendix 10: Results of the sensitivity analysis

Table S10: Results for the one-way sensitivity analysis

|  | **Median** | | | | | | | |
| --- | --- | --- | --- | --- | --- | --- | --- | --- |
|  | **Healthcare provider** | | | | **Societal** | | | |
|  | **KES** | **% difference** | **USD** | **% difference** | **KES** | **% difference** | **USD** | **% difference** |
| **Staff time (Adjusted)** | 32886.93 | 82.8% | 254.37 | 82.9% | 39478.03 | 64.5% | 305.37 | 64.5% |
| **Bed charges (Upper limit)** | 19038.17 | 5.8% | 147.27 | 5.9% | 25048.46 | 4.4% | 193.76 | 4.4% |
| **Base case** | 17988.17 | 0.0% | 139.11 | 0.0% | 23998.46 | 0.0% | 185.64 | 0.0% |
| **Bed charges (Lower limit)** | 16113.17 | -10.4% | 124.65 | -10.4% | 22123.46 | -7.8% | 171.13 | -7.8% |
|  | **Mean** | | | | | | | |
|  | **Healthcare provider** | | | | **Societal** | | | |
|  | **KES** | **% difference** | **USD** | **% difference** | **KES** | **% difference** | **USD** | **% difference** |
| **Staff time (Adjusted)** | 36013.61 | 79.6% | 278.54 | 79.6% | 42777.6 | 59.5% | 330.89 | 59.5% |
| **Bed charges (Upper limit)** | 21127.46 | 5.4% | 163.43 | 5.4% | 27891.45 | 4.0% | 215.74 | 4.0% |
| **Base case** | 20049.66 | 0.0% | 155.06 | 0.0% | 26813.65 | 0.0% | 207.41 | 0.0% |
| **Bed charges (Lower limit)** | 18125.01 | -9.6% | 140.21 | -9.6% | 24889 | -7.2% | 192.52 | -7.2% |

4	Kenya National Bureau of Statistics. The Kenya Poverty Report 2022. Nairobi, Kenya: Kenya National Bureau of Statistics (KNBS) 2024.

5	Kenya National Bureau of Statistics. *Kenya population and housing census*. Nairobi: Kenya National Bureau of Statistics 2019.

6	Kenya National Bureau of Statistics. *Kenya demographic and health survey, 2022*. Nairobi, Kenya: Kenya Nationa Bureau of Statistics 2023.

7	Kenya National Bureau of Statistics. *Providing, managing and promoting quality statistics (Economic Survery 2024)*. Nairobi, Kenya: Kenya National Bureau of Statistics 2024.

8	World Bank Group. Employment to population ratio, 15+, total (%) (modeled ILO estimate). World Bank Open Data. 2025. https://data.worldbank.org/indicator/SL.EMP.TOTL.SP.ZS (accessed 2 October 2025)
